# Development of a Novel Blood-Based Assay for Brain-Derived Tau and Its Validation in Traumatic Brain Injury

**DOI:** 10.64898/2026.06.05.26354965

**Authors:** Wasiu G. Balogun, Xuemei Zeng, Michel N. Nafash, Anuradha Sehrawat, Ruyu Shi, Sarah E. Svirsky, David O Okonkwo, Ava M. Puccio, Thomas K. Karikari

## Abstract

Brain-derived tau (BD-tau) is an emerging blood-based biomarker for neurodegeneration, yet there are currently limited well validated BD-tau assays available for research and clinical use. To enhance access to this vital biomarker for neurological disorders including traumatic brain injury (TBI), we developed a novel blood-based immunoassay for BD-tau on the ultra-sensitive Quanterix HD-X platform using Single Molecule Array technology. Analytical validation assessed dilution linearity, specificity, precision, detection limits, and spike recovery, each recording robust metrics in agreement with international expert recommendations. The assay demonstrated robust validation metrics, achieving between-run stability of 95% when analyzing aliquots from six independent plasma and serum samples across five analytical runs. It also showed strong dilution linearity when diluted four-fold and achieved over 90% recovery when spiked with cerebrospinal fluid. Next, we evaluated the clinical utility of the assay in cohorts of individuals with traumatic brain injury (TBI), where strong performances were recorded whether using the 2-step or 3-step assay formats (ρ= 0.94; p < 0.0001). Furthermore, plasma BD-tau distinguished samples from TBI patients based on time from injury and severity (AUC=0.93). Plasma BD-tau differentiated between favorable and unfavorable functional outcomes in the acute-severe group. Our findings underscore the significant potential of the BD-tau assay as a biomarker for TBI in the severe phase.

## 1 Introduction

Blood-based biomarkers (BBMs) are transforming our understanding of neurological disorders by providing cost-effective, accessible, and high-throughput assessments to monitor disease severity, outcomes, and progression patterns (Balogun et al., 2023; Schöll et al., 2024). These biomarkers have been applied to several neurological disorders, including traumatic brain injury (TBI). A notable development in BBMs for acute neurological disorders is the recent United States Food and Drugs Administration approval of a combined test panel of glial fibrillary acidic protein (GFAP) and ubiquitin carboxy-terminal hydrolase L1 (UCH-L1) as a biomarker for determining the need for computed tomography scan after mild TBI (Wichmann et al., 2025). While BBMs are valuable for enhancing our understanding of disease pathology, they can also play crucial roles in unravelling biological processes such as neuronal injury that may occur concurrently.

Neuronal injury, or neurodegeneration, is an important process present in several central nervous system disorders (Patel et al., 2025). In these conditions, neurons (and other cell types) are damaged or dysfunctional, leading to the release of specific cellular material that can disrupt the brain’s ability to transmit signals, resulting in diseases such as TBI, stroke, Alzheimer’s disease (AD), and multiple sclerosis (Brett et al., 2022; Patel et al., 2025). In neurological disorders characterized by acute injury, including TBI and acute ischemic stroke, current BBMs show measurable changes within hours of the impact. These include total tau (t-tau), neurofilament light chain (NfL), GFAP and UCH-L1 (Oris, Khatib-Chahidi, et al., 2024; Pattinson et al., 2020; Shahim et al., 2020). These biomarkers are highly useful as they capture different aspects of cellular injury. However, their presence in peripheral tissue introduces variability, as cerebrospinal fluid (CSF) measurements may differ from blood measurements due to peripheral contributions, particularly in the case of tau (Barthélemy et al., 2020; Fischer & Baas, 2020; Müller et al., 2017). To overcome these limitations, a CNS-specific biomarker called brain-derived tau (BD-tau) has been developed for measuring neuronal injury with high specificity in these disorders (Gonzalez-Ortiz, Turton, et al., 2023).

While BD-tau assays are now available from multiple sources, only two BD-tau assays have been well validated. The original method has been validated across dozens of independent studies related to acute neurological disorders such as TBI and ischemic stroke as well as chronic neurodegenerative diseases such as Alzheimer’s disease (Gonzalez-Ortiz, Dulewicz, et al., 2023; Gonzalez-Ortiz, Turton, et al., 2023; Gundersen et al., 2025). The second assay, developed by Quanterix Corporation, is a commercialized version of the original Gonzalez-Ortiz et al. method, and has undergone both analytical and clinical validation (Halbgebauer et al., 2026; Nafash et al., 2026; Zeng et al., 2026). In this context, we developed a novel BD-tau assay, Pitt-BD-tau. We then analytically validated it following established protocols recommended by the international consortium of clinical chemists (Andreasson et al., 2015) and clinically validated it using plasma samples from two TBI cohorts.

## 2 Methods

### 2.1 Plasma BD-tau immunoassay development

The development, validation, and clinical studies of the BD-tau assay were performed at the Biofluid Biomarker Laboratory, Department of Psychiatry, School of Medicine, University of Pittsburgh, Pittsburgh, PA, USA. An ultra-sensitive immunoassay was established using either a two (80-7 cadence) or three-step (80-7-7 cadence) format on the Single Molecule Array (Simoa)

HD-X platform (Quanterix, MA, USA). The Pitt-BD-tau assay was assembled with optimized concentrations and volumes of in-house, ready-to-use reagents, including biotinylation reaction buffer, bead wash buffer, bead conjugation buffer, and bead diluent. Additional reagents and consumables such as streptavidin-β-galactosidase (SβG) concentrate (lot no. 42470), SβG diluent (lot no. 424714), resorufin-β-d-galactopyranoside (RGP) (lot no. 319213), system wash buffer 1 (lot no. 20869400) and buffer 2 (lot no. 20908800), sealing oil (lot no. 20810700), 96-well plates (reference no. 103022), conductive tips (lot no. K195316I), array discs (lot no. 100001), cuvettes, and the HD-X instrument were procured from Quanterix, Inc. (Billerica, MA).

For the capture antibody, anti-human BD-tau monoclonal antibody (clone 2B8, mouse IgG1k) (lot. no. AK3569; Roboscreen, Leipzig, Germany) was coupled to the Quanterix paramagnetic beads (lot. no. 333608). According to the validation data provided by the supplier, the 2B8 antibody selectively binds to a continuous peptide sequence at the junction of exons 4 and 5 on CNS-abundant tau isoforms, specifically the six abundant isoforms expressed in the adult human brain (https://www.roboscreen.com/products/neurodegeneration/antibodies-for-tau-protein-and-beta-amyloid/). In addition, the 2B8 antibody does not recognize the protein epitope containing the exon 4a insert, including constructs spanning the contiguous exon 4–4a–5 region.

Biotin-conjugated mouse monoclonal antibody targeting the N-terminal region of tau (Tau12; BioLegend, cat. no. SIG-39416) was used for detection. Recombinant full-length tau441 (SignalChem, cat. no. T08-50FN-50) was employed as the assay calibrator. Plasma and serum samples were diluted with Quanterix Homebrew sample diluent B (lot no. 506221), while the Quanterix p-tau181 sample diluent (lot no. 526709) was used for both calibrator and detector dilutions.

### 2.2 Analytical validation

The following analytical validation parameters were examined: precision, lower limit of quantification (LLOQ), specificity, dilution linearity, and spike recovery, adhering to guidelines established by an international consortium of clinical chemists (Andreasson et al., 2015).

Precision was assessed using two plasma and four serum samples measured in five technical replicates across five different days, from which repeatability and intermediate precision were calculated. Analyte concentrations were normalized to the average of all 25 measurements for each respective sample and expressed as percentages.

LLOQ was determined from the four-parameter logistic (4PL) regression curve fitting after running 16 blank samples of the calibrator diluent, calculating the mean and standard deviation of the signal from a ten-point standard curve using recombinant tau441 (0–25 pg/mL; in duplicates). The LLOQ was estimated by extrapolating the signal corresponding to 10 standard deviations above the mean of blank measurements onto the calibration curve to obtain the corresponding concentration.

For specificity, we compared the Pitt-BD-tau assay signals generated by four different concentrations of recombinant tau441 relative to the same concentration of peripheral tau (a.k.a. big tau; sourced from Sino Biologicals US Inc.). Tau441 and big tau, at concentrations ranging from 3.125 to 50 µg/mL, were spiked into plasma samples (QCA), followed by 4X dilutions with the sample diluent, and subsequently analyzed using the Pitt BD tau assay.

Dilution linearity was evaluated using three plasma samples, each diluted four-fold with sample diluent and divided into seven aliquots. A calibrator stock solution (750 pg/mL) was prepared and serially diluted multiple times in pooled plasma sample (to enable assessment of potential matrix effect) to generate the following theoretical concentrations below the analytical LLOQ (pg/mL): 93.75, 23.44, 5.859, 1.465, 0.3662, and 0.0916. To assess relative errors, concentrations were normalized to the mean of the undiluted sample and corrected for dilution by multiplying by the respective dilution factors. Results in the range of 80-120% of the original signal after adjustment were considered as acceptable linear dilution.

Spike recovery was determined by spiking CSF with known BD-tau concentration into both serum sample and assay diluent. Recovery was calculated as the difference in signal between spiked and un-spiked serum, relative to the corresponding difference observed when spiked into the assay diluent.

### 2.3 Study participants

This study involved control, chronic-mixed and acute-severe TBI participants. The chronic-mixed TBI and control individuals were recruited through a Peer Reviewed Alzheimer’s Research Program (PRARP) study. Inclusion criteria for the chronic-mixed TBI group were: 1. A history of at least one TBI occurring more than one year before enrollment.; 2. Age 29-59 years; 3. Suspected cognitive impairment which was based on neuropsychological assessment or professional diagnosis of cognitive impairment. Cognitive status was assessed using three questions from the Rivermead Post-Concussion Symptom Questionnaire (i.e., forgetfulness/poor memory; poor concentration; taking longer to think). At enrolment, all participants had Glasgow coma scale (GCS) score of 15. The exclusion criteria were: 1. History of penetrating TBI, 2. history of pre-existing neurologic or neurodegenerative disorder, 3. History of psychiatric disorder, 4. Active drug or alcohol dependence, 5. Contraindication to magnetic resonance imaging or PET scans. For the control group, the inclusion criteria were age 29-59 years, while the exclusion criteria included a history of blast exposure or moderate to severe TBI.

The severe TBI participants were prospectively enrolled at admission between 2019-2024 through the Brain-Trauma Research Center (BTRC) at the University of Pittsburgh, Pittsburgh, PA, USA. Eligibility was determined by the presence of severe TBI defined as GCS score <9 at enrollment and age 16–80 years. Exclusion criteria were penetrating brain injury, brain death and pregnancy.

### 2.4 Ethical Approval

The study was approved by the Institutional Review Board at the University of Pittsburgh (19110161 for PRARP and 19030228 for BTRC). Written informed consent was obtained from the participants for the PRARP study and by the patients’ legal authorized representative for the BTRC study. This study adhered to the Strengthening of the Reporting of Observational Studies in Epidemiology (STROBE) reporting guidelines.

### 2.5 Clinical assessment of TBI severity and outcome

In the acute-severe TBI group, injury severity was clinically assessed using the Glasgow Coma Scale (GCS) during study enrollment and functional outcome was assessed by the Glasgow Outcome Scale Extended (GOS-E) at 6 months. For the GOS-E, a score between 1 and 4 indicated an unfavorable outcome, while a score between 5 and 8 indicated a favorable outcome. Outcome assessments were collected using a mixed-methods approach, including interviews conducted in person or via telephone. For participants with substantial impairment, their proxies were interviewed. The GOS-E score was also used for survivability, 1 was considered as death while score between 2 and 8 indicated survival.

### 2.6 Blood sample handling and biomarker measurements

Plasma samples were collected from all 63 participants. Up to 15ml of blood was collected per patient at one time. Plasma was collected with a 6ml K2EDTA tube (BD Medical, cat no. 367863). Plasma was processed using EDTA tube by centrifugation at 2,000g for 10 min and aliquoted and stored at-80 freezer until used. Chronic-mixed and control blood was taken at baseline. Acute-severe blood samples were taken four days post-injury.

For the validation experiments, only CSF from the control subjects were used in the study. There were 4 participants whose CSF was collected via lumbar puncture (lumbar region). 10 mL of CSF was collected from each participant. Immediately after collection, CSF was gently inverted to avoid possible gradient effects, briefly centrifuged at low speed, pooled together and aliquoted into polypropylene cryovials prior to freezing and storage.

All samples were processed according to standard procedures and stored at-80 °C until use (Zeng et al., 2024). For the commercial BD-tau, BD-tau Adv PLUS reagent kit (Quanterix, lot no. 504274) was used. The Pitt-BD-tau levels were measured on the Simoa HD-X platform using both the 2-step and 3-step protocols for comparison.

### 2.7 Statistical analysis

Statistical analyses were conducted using Prism version 9.3.1 (GraphPad Software) and R version 4.4.0. No priori sample size calculation was performed for this study. The sample size was determined by the availability of well-characterized plasma samples within the cohort, reflecting the exploratory and proof-of-concept nature of this work. This approach is consistent with prior biomarker studies in traumatic brain injury (Gonzalez-Ortiz, Dulewicz, et al., 2023; Okonkwo et al., 2013), in which sample sizes are often constrained by cohort availability and biospecimen access.

The normality of data distributions was assessed with the Kolmogorov-Smirnov test. Since the data were not normally distributed, nonparametric tests were employed, and continuous data are reported as median (IQR) values. Spearman correlation was used to examine concordance between the 2-step and 3-step assay protocols. Outlier testing was performed for the Spearman correlation analysis between 2-step and 3-step plasma BD-tau measurements using studentized residuals and Cook’s distance derived from a linear regression model. To compare plasma BD-tau levels between TBI clinical outcome groups (unfavorable and favorable outcomes), mean fold differences (95% CI) were calculated, and statistical comparisons were made using the Mann-Whitney U test. Differences in biomarker levels across the entire cohort or within each outcome group were evaluated using the Kruskal–Wallis rank sum test, with Dunn’s post hoc comparisons and adjustment for multiple testing. Two sided p-values below 0.05 were considered statistically significant. Classification analyses were performed using logistic regression models adjusted for age and sex. Separate pairwise binary models were fit to discriminate severe-acute cases from controls and from chronic-mixed cases. Model discrimination was quantified using the area under the receiver-operating characteristic curve (AUC) based on predicted probabilities. To address the limited sample size and reduce potential overfitting, model performance was assessed using stratified k-fold validation. Specifically, stratified 5-fold cross-validation was repeated 1,000 times with different random fold assessments, and AUCs were calculated from out-of-fold predicted probabilities. Empirical 95% confidence intervals for the AUC were obtained from the distribution of cross-validated estimates across repeats. Probability thresholds were selected within each training fold using Youden’s index and applied only to the corresponding held-out test data. Threshold-dependent performance metrics, including sensitivity, specificity, accuracy, positive predictive value, and negative predictive value, were summarized across cross-validation repeats using the median and interquartile range.

## 3 Results

### 3.1 Pitt-BD-tau assay development

The assay development workflow for the ultrasensitive detection of Pitt-BD-tau using the Simoa platform is shown in Figure 1A illustrating both the 2- and 3-step assay formats.

**Figure 1:**
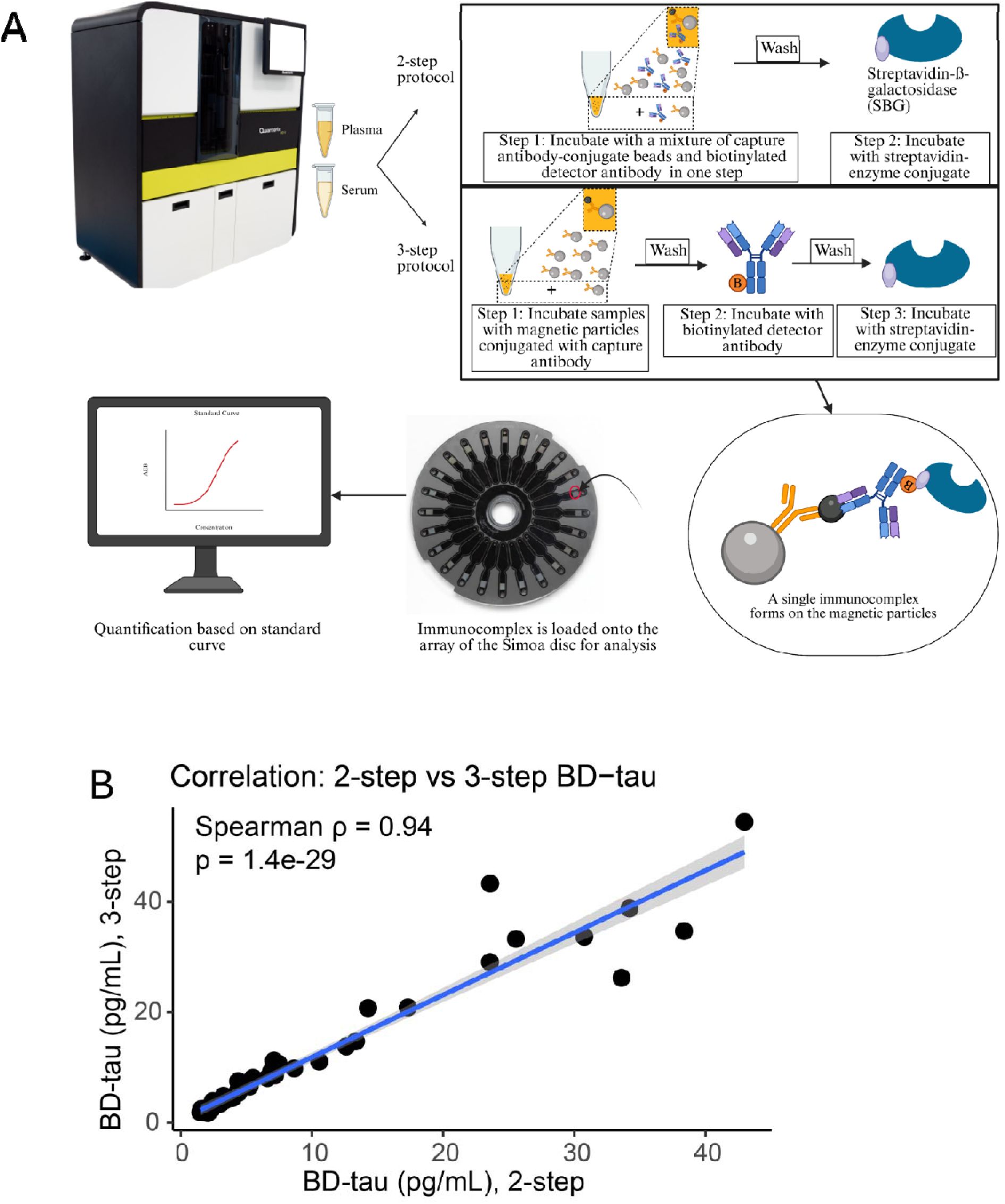
Workflow of the plasma Pitt-BD-tau assay development. (A). Schematic illustration of the workflow of Pitt-BD-tau assay development using the ultrasensitive Simoa HDX platform. The 2-step and 3-step homebrew configurations were adopted. In the 2-step assay, the sample is first incubated with the paramagnetic bead-coated capture antibody and detector antibody, followed by the addition of SβG. While, the 3-step assay involves an initial incubation with the paramagnetic bead-coated capture antibody, followed by the detector antibody, and finally the addition of SβG. **(B).** Spearman rank correlation between Pitt-BD-tau concentrations measured in identical clinical samples using either the 2-step or the 3-step assay formats. Each point represents an individual sample. The solid line indicates the fitted linear regression used for visualization, and the grey shaded area represents the 95% confidence interval around the regression line.

Assay optimization was performed using both approaches; however, the 3-step protocol was ultimately selected as it demonstrated superior performance, including an improved detection range and lower quantitation limit. The 2-step and 3-step methods are analogous to the sandwich direct and indirect enzyme-linked immunosorbent assay (ELISA) procedures, respectively (Thangavelu & Boutté, 2021). In the 2-step format, the sample is first incubated with the paramagnetic bead-coated capture antibody and detector antibody simultaneously, followed by a wash and addition of SβG. In contrast, the 3-step format involves sequential incubations: first with the paramagnetic bead-coated capture antibody, followed by a wash, then with the detector antibody, another wash, and finally the addition of SβG (Thangavelu & Boutté, 2021).

### Correlation between the 2-step and 3-step assay formats

We applied both the 2-step and 3-step assays to the same set of clinical (plasma or serum) samples. The results showed a strong Spearman correlation between two protocols (ρ = 0.94, p = 1.4 × 10□²□), suggesting that the assay has high consistency regardless of protocol (Figure 1B).

### 3.2 Analytical validation

After antibody selection and optimizing the assay steps, buffers and reagent concentrations, analytical validation was conducted using both plasma and serum samples. The Pitt-BD-tau assay was calibrated with a ten-point standard curve, consisting of nine non-blank calibrators ranging from 0.103 pg/mL to 25 pg/mL. A representative calibration curve is shown in Supplemental Figure 1.

The AEB signals for the 16 blank samples (Supplemental Table 1) ranged from 0.116 to 0.133 with the mean and SD being 0.125 and 5.11×10^-3^ respectively. The LLOQ signal (mean + 10 SD) gives a value of 0.177, which corresponds to an analytical LLOQ of 0.012 pg/mL and functional LLOQ of 0.047 pg/ml (after accounting for 4X sample dilution).

To determine if the assay is specific for the CNS-abundant tau isoforms, we compared Pitt-BD-tau signals in equal concentrations (3.125 pg/ml, 12.5 pg/ml, 25 pg/ml and 50 pg/ml) of recombinant versions of the CNS-abundant 2N4R (tau-441) vs. the peripheral-abundant big-tau isoforms. The assay exclusively recognized the tau-441 isoform in a concentration-dependent manner but not big tau. The big tau values were comparable to the blank values indicating that the Pitt-BD-tau assay was specific for CNS-abundant tau isoforms (Figure 2A). Collectively, these results demonstrate that the Pitt-BD-tau assay specifically quantifies BD-tau and does not cross-react with big tau.

**Figure 2:**
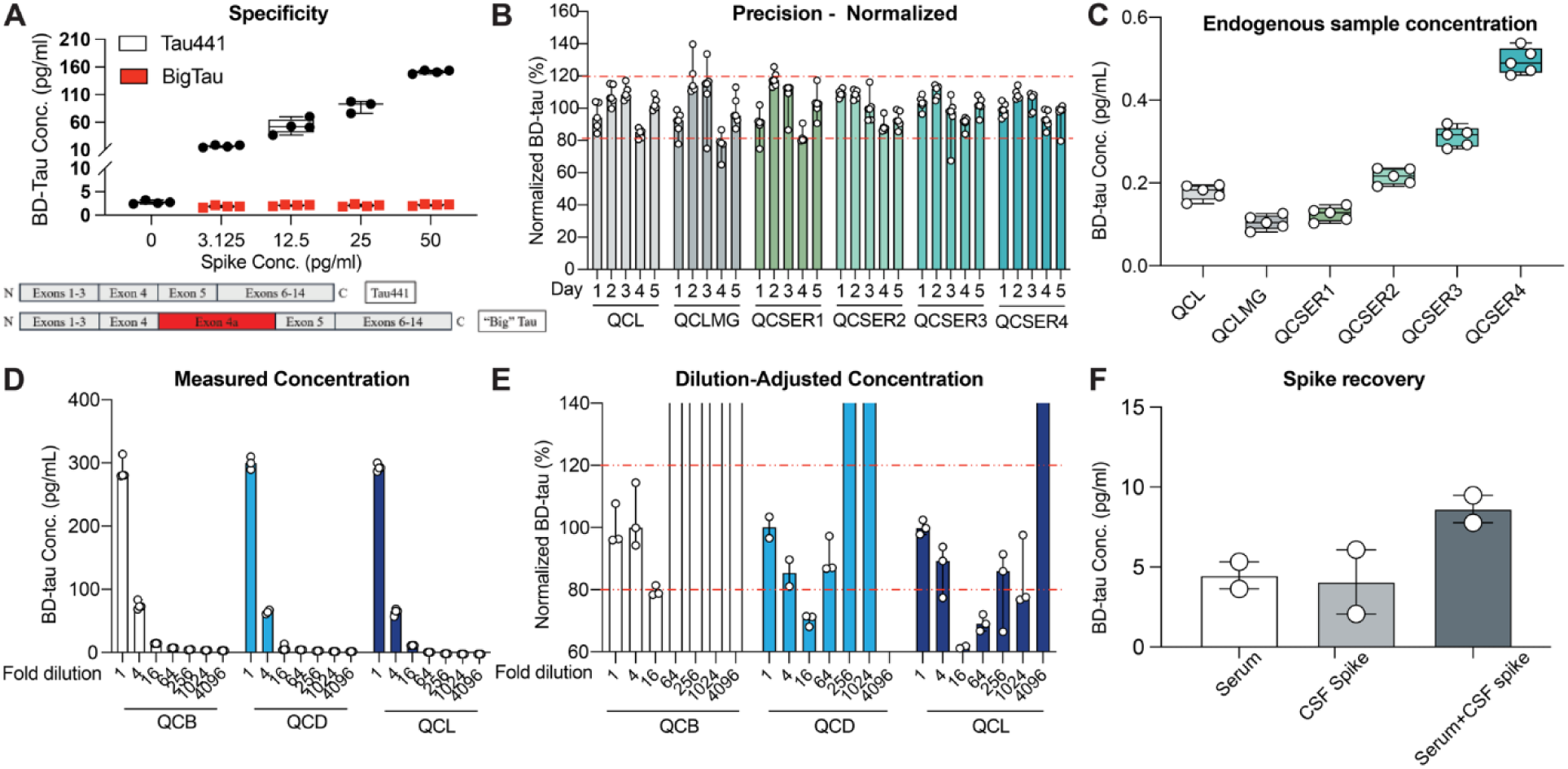
Analytical validation of the plasma Pitt-BD-tau assay. Unless otherwise indicated, points represent individual repeated measurements (or run-level measures where applicable), with the number of points corresponding to the sample size. Boxes or bars represent the median, and error bars represent the interquartile range (IQR; 25^th^-75^th^ percentile), and box whiskers extend to values within 1.5 times of the IQR. **(A) Specificity.** Comparison of the BD-tau assay readings when measured in equal molar concentrations of recombinant versions of CNS-abundant tau (2N4R isoform) and the big tau isoforms at 0, 3.125, 12.5, 25, and 50 pg/ml spiked into an identical plasma sample. **(B) Precision.** Normalized plasma BD-tau concentrations for the six samples evaluated in the precision experiments. Single-use aliquots of each specimen were measured 5 individual times, across five runs over five different days. **(C)** Unnormalized BD-tau concentrations for the same six specimens. **(D) Dilution linearity.** Portrayal of dilution linearity of plasma samples showing the concentration from non-diluted (ND) up to 4096-fold diluted, across four-fold stepwise decreases. **(E)** The dilution-adjusted relative errors for samples showing the fold dilutions the adjustment of which produced concentration within ±20% the normalized concentration of the dilution-free (neat) sample (shown as dotted red lines at 80% and 120%). **(F) Spike recovery.** Spike recovery in a serum sample spiked with CSF. Bars show the group median concentration (pg/mL).

For the precision test, we measured four serum and two plasma samples across five different analytical days, with 5 replicates per sample on each day. Figure 2B illustrates the relative distribution of measurements, normalized to the mean value of all measurements for each corresponding sample, across the five days. The measured concentrations of each sample are shown in Figure 2C. Intra-plate CV and inter-plate coefficients of variation (%CVs) are provided in Supplemental Table 2. The assay demonstrated intra-plate CVs ranging from 5.14% to 12.17% and inter-plate CVs ranging from 8.04% to 19.01%, reflecting intermediate precision and repeatability within the acceptable limits defined by the international consortium of clinical chemists.

For dilution linearity, three unique plasma samples were 4X serially diluted with the assay diluent from 1 to 4096 (Figure 2D). Among the dilutions tested, only the 4-fold dilution produced an optimal signal with dilution-adjusted concentration between 80% to 120% of the undiluted value in all the plasma samples (Figure 2E). The results show that the Pitt-BD-tau assay gives optimal signals when samples are diluted up to 4-fold.

Spike recovery was evaluated by spiking a neat CSF sample into four-fold diluted serum samples. The assay demonstrated a recovery of 97% of the expected analyte signal (Figure 2F) which indicates a lack of matrix interference and a high degree of accuracy.

### 3.3 Clinical validation in the TBI cohorts

#### Demographic information

A total of 63 participants provided plasma samples (Table 1). In the TBI group (n=55), 21 were chronic-mixed while 34 were acute-severe. In the chronic-mixed group, 81.0% of participants were male, with a median age of 40 years while the acute-severe comprised of older participant with median age of 51 years, 77.7% were male. In contrast, the control group consisted of 87.5% male participants, with a median age of 40 years. The most common cause of injury was motor vehicle accidents (51.9%), while the least common cause was assault/fight-related injury (5.6%).

### Association with disease severity

#### Association between BD-tau levels and injury severity as measured by Pitt-BD-tau assay

To examine the association between BD-tau and injury severity, plasma BD-tau concentrations were compared across clinical groups and subgroups using both 2-step and 3-step Pitt-BD-tau assay protocols.

Wilcoxon rank-sum showed no difference in Pitt-BD-tau readings between the control and the chronic-mixed groups, however the comparisons between the control and acute-severe and chronic-mixed vs acute-severe were highly significant (p < 0.0001), for both assay formats. Figures 3A and 3B present box plots illustrating the distribution of Pitt-BD-tau concentrations across different levels of TBI severity: control, chronic-mixed, and acute-severe. An overall Kruskal–Wallis test confirmed highly significant group effects (two-step: p = 1.30 × 10 ¹; three-step: p = 4.74 × 10 ¹). The full statistical reports are provided in Supplementary Table 3.

**Figure 3:**
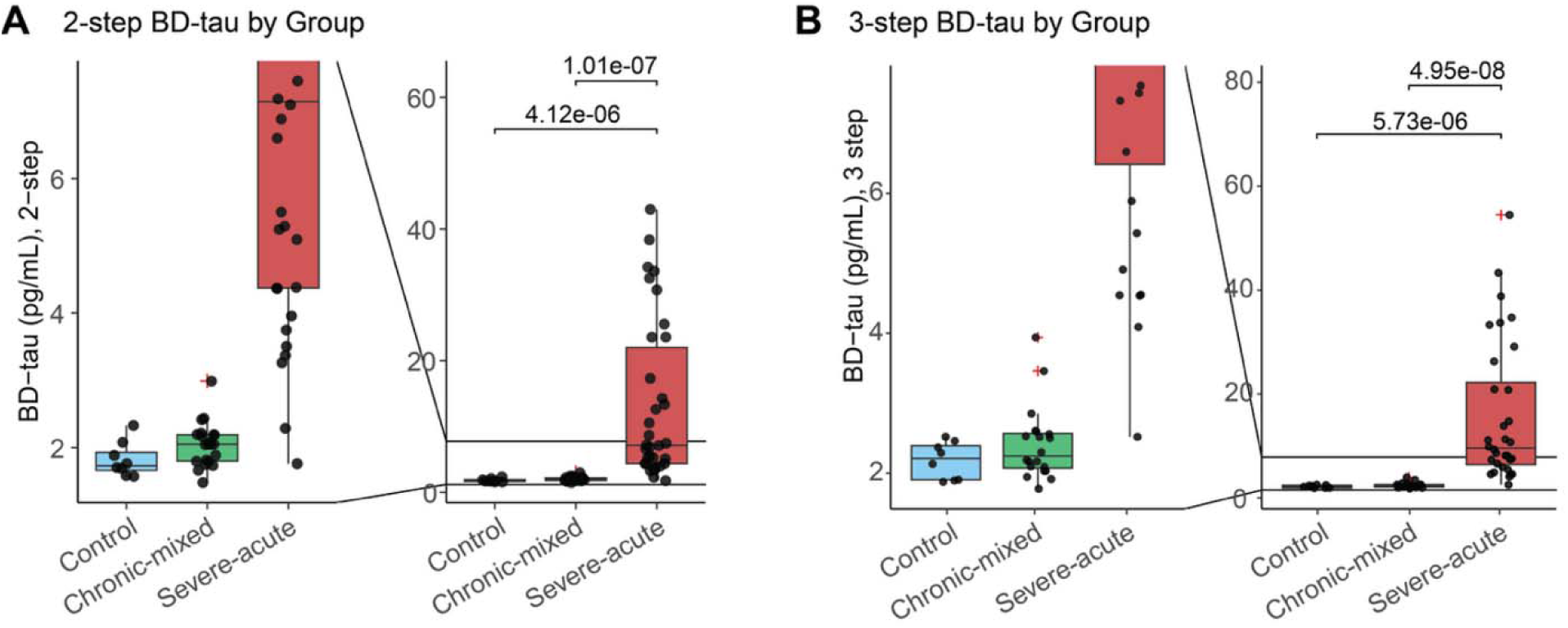
Association of BD-tau with TBI severity. Unless otherwise indicated, each point represents an individual sample, with the total number of points corresponding to the sample size. Boxes display the median and interquartile range (IQR; 25^th^-75^th^ percentile), and whiskers extend to values within 1.5 times of the IQR. Group differences were assessed using the Kruskal-Wallis test. Following a significant overall effect, pairwise group comparisons were performed using Dunn’s post-hoc test with Bonferroni correction. Annotated p-values represent Bonferroni-adjusted p-values from these pairwise comparisons. **A.** Box plots showing the distribution of plasma BD-tau concentrations for control, chronic-mixed and acute-severe in the 2-step assay protocol. Controls N = 8; Chronic-Mixed N = 21; Severe-acute N = 33. **B.** Box plots showing th distribution of the BD-tau for control, chronic-mixed and acute-severe in the 3-step assay protocol. Controls N = 8; Chronic-Mixed N = 20; Severe-acute N = 33.

### Association with clinical outcomes

#### Pitt-BD-tau assay and functional outcome

To evaluate prognostic value, TBI participants in the acute-severe group were stratified into favorable and unfavorable outcome groups. The assay effectively distinguished between these outcome groups, as indicated by the significantly higher plasma BD-tau levels in the unfavorable vs. favorable outcome groups (p<0.05) using both 2-step (Figure 4A) and 3-step (Figure 4C) assay formats.

**Figure 4:**
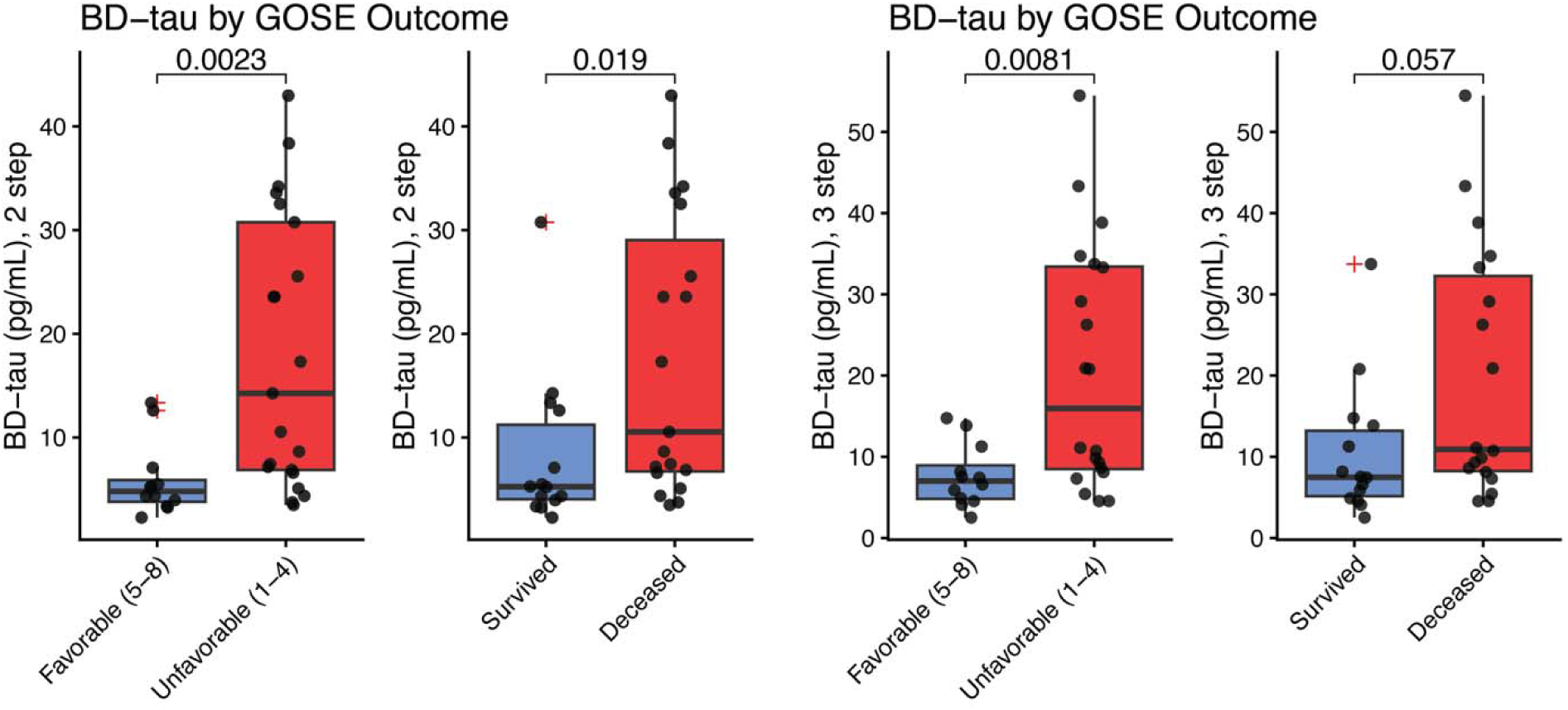
Association of plasma BD-tau with functional outcome after TBI. Unless otherwise indicated, each point represents an individual sample, with the total number of points corresponding to the sample size. Boxes display the median and interquartile range (IQR; 25^th^-75^th^ percentile), and whiskers extend to values within 1.5 times of the IQR. Annotated p-values represent two-sided p-values from pairwise Wilcoxon rank-sum tests. **A.** Levels of Pitt-BD-tau on day four after injuries indicated a significant difference between favorable and unfavorable outcomes in the 2-step protocol. Favorable (5-8) N = 12; Unfavorable (1-4) N = 21. **B.** Levels of Pitt-BD-tau on day four after injuries to predict survival or death in the 2-step protocol. Survived N = 14; Deceased N = 18. **C.** Levels of the Pitt-BD-tau assay measured at four days post-injury showed a significant difference between favorable and unfavorable outcomes in the 3-step assay protocol. Favorable (5-8) N = 12; Unfavorable (1-4) N = 20. **D.** Levels of Pitt-BD-tau on day four after injuries to predict survival or death in the 3-step protocol. Survived N = 14; Deceased N = 19.

#### Pitt-BD-tau assay and survivability

To further evaluate if the assay can predict if a TBI participant will survive, the acute-severe group was stratified into deceased and survived outcome groups. The assay was able to distinguish between these groups in the 2-step (Figure 4B) but not in the 3-step (Figure 4D) assay formats.

#### Preliminary values for Pitt-BD-tau level as a potential outcome predictive test for TBI

Table 2 presents the cross-validated classification performance of age-and sex-adjusted logistic regression models for discriminating severe-acute cases from controls and from chronic-mixed cases. All three plasma biomarkers demonstrated strong discriminative ability for distinguishing severe-acute cases from controls. Cross-validated AUCs were consistently high for both the 2-step and 3-step Pitt-BD-tau assays (AUC = 0.968-0.972), with empirical 95% confidence intervals spanning approximately 0.935 to 0.996, indicating robust separation between groups. Similarly, discrimination between severe-acute and chronic-mixed cases remained high across biomarkers, with cross-validated AUCs ranging from approximately 0.931 to 0.937 and corresponding confidence intervals of approximately 0.875 to 0.970. Threshold-dependent performance metrics, including sensitivity, specificity, accuracy, positive predictive value, and negative predictive value, were estimated using probability thresholds selected within training folds and summarized across cross-validation repeats using the median and interquartile range. The optimal probability thresholds were stable across cross-validation runs, supporting the robustness of the observed classification performance. The unadjusted model of the performance metrics is presented in Supplementary Table 4.

## 4 Discussion

A major challenge in clinical practice with the use of blood-based t-tau is the discordance between t-tau levels in plasma and serum samples vs. CSF, as approximately 80% of the circulating t-tau signal is thought to originate from peripheral sources (Barthélemy et al., 2020). This discrepancy can lead to inaccurate assessments of functional outcomes and the severity of acute neurological injury including TBI. In contrast, as shown previously, BD-tau has much stronger concordance between CSF and plasma levels (Nafash et al., 2026). The Pitt-BD-tau biomarker described here selectively measured tau derived from the brain, making it suitable for quantifying the true extent of neurological injury using accessible blood samples. Implementing the newly developed Pitt-BD-tau assay in clinical settings could improve the accuracy and speed of disease outcome assessments, providing clinicians with a reliable biomarker that can be measured shortly after an injury, thus facilitating more precise treatment planning. The Pitt-BD-tau assay offers a comparative advantage due to its affordability. While the earlier assay utilized sheep monoclonal antibodies (Gonzalez-Ortiz, Turton, et al., 2023), Pitt-BD-tau was developed with mouse monoclonal antibodies, a more cost-effective and widely accessible alternative.

The assay demonstrated strong sensitivity, exhibiting a four-fold dilution linearity comparable to findings from similar studies (Gonzalez-Ortiz, Turton, et al., 2023; Nafash et al., 2026). Additionally, it showed robust stability in both between-run and within-run tests for serum and plasma samples, confirming its utility in human specimens. The low value observed in the lower limit of quantification (LLOQ) test indicates the assay’s capability to accurately detect target molecules at low concentrations, thereby broadening its applicability in various clinical settings and potentially streamlining the diagnostic process. Furthermore, the assay achieved approximately 100% recovery after spiking serum with cerebrospinal fluid (CSF), underscoring its high accuracy based on trueness of measured values.

The GOS-E and the GCS are recognized as gold standard tools for assessing functional outcomes and the severity of TBI (Chawla et al., 2020; Okonkwo et al., 2013; Wilson et al., 1998). While the original GOS is widely used, GOS-E is considered more sensitive to changes in patient status (McMillan et al., 2016; Wilson et al., 1998). The GCS is particularly useful for assessing consciousness in patients with severe traumatic injuries, helping to prevent premature discharges (Mattei & Teasdale, 2020; Nell et al., 2000). Both tools were utilized in this study, as they are used in the clinic for evaluating patient recovery and tailoring rehabilitation plans. They provide critical data that enable healthcare professionals to make informed treatment decisions. Moreover, their standardized nature allows for consistent evaluations across different clinical settings, enhancing the comparability of research findings (Mattei & Teasdale, 2020). The Pitt-BD-tau was able to discriminate severe TBI from a milder TBI injury, that the chronic-mixed group represents (figure 3) as well as discriminate poor and favorable long-term outcomes (figure 4). This showed Pitt-BD-tau showed strong correlation with these widely used clinical outcomes scoring systems for TBI, highlighting its high potential as a predictive biomarker of TBI severity.

Nonetheless, the TBI outcome scores do not account for the neurochemical processes occurring in injured patients, such as neuronal damage (Okonkwo et al., 2013). This is where the Pitt-BD-tau assay demonstrates significance as it can measure biochemical changes that are reflective of functional outcome and trauma severity. Previous studies have shown the potential of BD-tau in this area (Gonzalez-Ortiz, Dulewicz, et al., 2023; Hicks et al., 2025). The outcomes for all participants in the chronic TBI group were favorable (Table 1), eliminating the need for further analysis. In contrast, the results for the acute-severe TBI group indicated that the BD-tau assay could predict functional outcomes as early as four days post-injury in both assay formats.

Additionally, the study explored whether the BD-tau assay could distinguish between those who died from severe injury and those who survived; the assay was able to discriminate between the survived and deceased groups. Interestingly, the assay was only able to significantly differentiate between these groups in the 2-step assay format. Meanwhile, the difference in significance between the 2-step and 3-step assays in predicting the survival outcome likely reflect differences in how each format preserves variability in BD-tau levels within clinically homogeneous subgroups. After restricting the analysis to the severe-acute group, residual variability in injury severity and biomarker concentrations is limited. Under these conditions, the 2-step assay appears to retain greater dispersion at higher BD-tau concentrations, resulting in larger rank separation between survival outcome groups and a statistically significant Wilcoxon test. In contrast, the 3-step assay, which is optimized for analytical specificity and precision, may partially compress the upper range of BD-tau values, thereby attenuating rank differences relevant for survival prediction in an unadjusted analysis. Further analysis in a larger cohort is needed to determine this capability.

To establish that the Pitt BD-tau was similar to other BD-tau assays, we compared the Pitt BD-tau with the Quanterix (commercial) BD-tau. Our results were comparable to the Quanterix BD-tau. Across all pairwise comparisons, the discriminative performance of the Pitt BD-tau assays was comparable to that of the Quanterix assay, with overlapping cross-validated AUCs. For example, in distinguishing severe-acute TBI cases from controls, the mean cross-validated AUCs were approximately 0.97 for both the commercial assay and the Pitt BD-tau assays. Similarly, in distinguishing severe-acute from chronic-mixed cases, cross-validated AUCs ranged from approximately 0.93 to 0.94 across all assays. These results indicate that, within this cohort, the Pitt BD-tau assays demonstrate classification performance comparable to that of the commercial BD-tau assay. We note, however, that the present study was not designed as a formal analytical head-to-head comparison (e.g., assessment of assay precision, calibration, or limits of detection). Therefore, while discriminative performance was similar, definitive conclusions regarding analytical superiority will require dedicated comparative validation studies.

A key strength of this study is the consistency of Pitt-BD-tau measurements across both the 2-step and 3-step assay formats, underscoring the robustness and reliability of the assay. However, based on results here the 3-step assay format seems more appropriate to be selected for future study.

Another important finding from this study is the optimal cutpoint strategy we developed to distinguish between control individuals and those with TBI, as well as to differentiate chronic TBI from acute TBI. This finding is significant given the challenges in classifying TBI based on disease severity. Interestingly, the adjustment for age and sex did not materially change the high performance of the BD-tau. From the unadjusted ROC, we further established a preliminary threshold of 2.80 and 3.31 pg/ml for separating severe-acute TBI from controls for the 2-step and 3-step assay respectively while 3.13 and 4.02 pg/ml for distinguishing severe TBI from chronic TBI in the 2-step and 3-step assay respectively. To our knowledge, this is the first study to implement a cutoff strategy for BD-tau in TBI, similar to established thresholds for other biomarkers (Hier et al., 2021; Okonkwo et al., 2013; Oris, Kahouadji, et al., 2024). Meanwhile, the unadjusted ROC analysis demonstrated near-perfect discriminatory performance of the biomarker. After adjustment for age and sex, the AUC remained high, ranging approximately from 0.93 to 0.97 across cross-validation repetitions. Although the sample size is modest, we used repeated cross-validation to mitigate overfitting and improve the internal validity of these estimates. Nevertheless, further adjustment for additional covariates and external validation in independent cohorts will be necessary.

This study does have its limitations, including (1) the lack of neuroimaging data, such as computed tomography scans, and neuropsychological assessments, (2) a predominantly male small sample size, and (3) reliance on retrospective self-report measures assessed at a single time point which makes the classification of participants in the chronic-mixed difficult. Long-term follow-up studies are necessary to address these limitations and provide further insights into the trajectories of symptom severity and Pitt-BD-tau concentrations over time. Future research should also consider covariate factors that may influence Pitt-BD-tau and neuropsychological outcomes, such as genetic predisposition, physical activity, and other lifestyle variables. Nonetheless, our findings suggest that Pitt-BD-tau is a promising candidate biomarker that may enhance our understanding of the persistent psychological symptoms experienced by individuals with TBI, particularly in cases of chronic and severe injuries. Finally, standardizing the assay process across different laboratories could further improve reliability and reproducibility.

## 5 Conclusion

We have successfully developed a BD-tau assay that demonstrates high specificity and strong recovery performance and the promise to differentiate both severity and outcome in TBI. Beyond TBI, this assay holds promise for broader applications in various other neurological disorders such as AD and ischemic stroke, which are currently explored in our lab.

## Author Contributions

**Wasiu Balogun:** conceptualization, investigation, visualization, formal analysis and writing – review and editing.

**Michel Nafash:** investigation, visualization, formal analysis, writing – review and editing.

**Anuradha Sehrawat:** formal analysis and writing – review and editing.

**Xuemei Zeng:** conceptualization, investigation, formal analysis, supervision, writing – review and editing.

**Ruyu Shi:** conceptualization, formal analysis, writing – review and editing.

**Sarah Svirsky:** resources, formal analysis and editing

**David O Okonkwo:** conceptualization, funding acquisition, supervision, resources and editing.

**Ava Puccio:** conceptualization, funding acquisition, supervision, resources and editing.

**Thomas Karikari:** conceptualization, investigation, funding acquisition, supervision, writing – review and editing.

## Data Availability

All data produced in the present study are available upon reasonable request to the corresponding authors

## Acknowledgements

We thank study participants, their families and caregivers for their participation in this cohort study.

## Data Availability Statement

The datasets used and analyzed in the current study are available from the corresponding authors upon reasonable request for the sole purpose of replicating the results in this study, provided a data use/sharing agreement is established in agreement with IRB stipulations and applicable laws in the Commonwealth of Pennsylvania and the United States.

## Abbreviations

AD: Alzheimer’s disease
AUC: Area under the curve
BBMs: Blood-based biomarkers
BD-tau: Brain derived-tau
CNS: Central nervous system
CSF: Cerebrospinal fluid
%CV: Coefficient of Variation
ELISA: enzyme-linked immunosorbent assay
FDA: Food and Drug Administration
GCS: Glasgow Coma Scale
GOS-E: Glasgow Outcome Scale Extended
LLOQ: Lower limit of quantification
Simoa: Single molecule array
TBI: Traumatic brain injury
t-tau: Total tau

