## Supplementary data for "Development of a Novel Blood-Based Assay for Brain-Derived Tau and Its Validation in Traumatic Brain Injury"

**Supplementary Tables**

**Table 1**. LLOQ AEB values

| Replicate | AEB value |
| --- | --- |
| LLOQ1 | 0.127457 |
| LLOQ2 | 0.132806 |
| LLOQ3 | 0.127172 |
| LLOQ4 | 0.123612 |
| LLOQ5 | 0.12205 |
| LLOQ6 | 0.120689 |
| LLOQ7 | 0.123146 |
| LLOQ8 | 0.129976 |
| LLOQ9 | 0.132702 |
| LLOQ10 | 0.132628 |
| LLOQ11 | 0.130304 |
| LLOQ12 | 0.127551 |
| LLOQ13 | 0.119809 |
| LLOQ14 | 0.116182 |
| LLOQ15 | 0.121315 |
| LLOQ16 | 0.120281 |

**Table 2.** Intra- and inter-plate coefficients of variation

| Sample | Intra-plate CV (%) | Inter-plate CV (%) |
| --- | --- | --- |
| QCL | 5.482 | 10.992 |
| QCLMG | 12.177 | 19.010 |
| QCSERUM1 | 8.180 | 15.052 |
| QCSERUM2 | 5.147 | 9.679 |
| QCSERUM3 | 6.993 | 10.262 |
| QCSERUM4 | 5.964 | 8.039 |

**Table 3.** Full statistical report on TBI severity, outcome and survivability

| **Figure 3A** | **Group (Control, Chronic-mixed, and Severe acute) differences in 2-step BD-tau** | | | | | |
| --- | --- | --- | --- | --- | --- | --- |
|  | **Kruskal-Wallis** H = 42.518, df = 2, p = 5.85E-10 | | | | | |
|  | **Dunn's post-hoc** | **group1** | **group2** | **z** | **p_raw** | **p_bonf** |
|  |  | Control | Chronic-mixed | 0.879 | 0.380 | 1.000 |
|  |  | Control | Severe-acute | 4.830 | 1.37E-06 | 4.12E-06 |
|  |  | Chronic-mixed | Severe-acute | 5.520 | 3.36E-08 | 1.01E-07 |
|  | **ANCOVA adjusted for sex and age (using log10-transformation on biomarkers):**  F(2,57) = 45.02, p = 1.86E-12 | | | | | |
|  | contrast | estimate | SE | df | t.ratio | p.value |
|  | Control - (Chronic-mixed) | -0.041 | 0.118 | 57 | -0.346 | 0.730 |
|  | Control - (Severe-acute) | -0.711 | 0.113 | 57 | -6.306 | 6.77E-08 |
|  | (Chronic-mixed) - (Severe-acute) | -0.670 | 0.080 | 57 | -8.379 | 4.85E-11 |
| **Figure 3B** | **Group (Control, Chronic-mixed, and Severe acute) differences in 3-step BD-tau** | | | | | |
|  | **Kruskal-Wallis** H = 42.940, df = 2, p = 4.74E-10 | | | | | |
|  | **Dunn's post-hoc** | **group1** | **group2** | **z** | **p_raw** | **p_bonf** |
|  |  | Control | Chronic-mixed | 0.654 | 0.513 | 1.000 |
|  |  | Control | Severe-acute | 4.760 | 1.91E-06 | 5.73E-06 |
|  |  | Chronic-mixed | Severe-acute | 5.650 | 1.65E-08 | 4.95E-08 |
|  | **ANCOVA adjusted for sex and age (using log10-transformation on biomarkers):**  F(2,55) = 51.5, p = 2.49E-13 | | | | | |
|  | contrast | estimate | SE | df | t.ratio | p.value |
|  | Control - (Chronic-mixed) | -0.027 | 0.111 | 55 | -0.245 | 0.808 |
|  | Control - (Severe-acute) | -0.706 | 0.106 | 55 | -6.654 | 2.08E-08 |
|  | (Chronic-mixed) - (Severe-acute) | -0.679 | 0.077 | 55 | -8.826 | 1.20E-11 |
| **Figure 4A** | **GOSE Outcome (Favorable 5-8 vs. Unfavorable 1-4) differences in 2-step BD-tau** | | | | | |
|  | **Two-sided Wilcoxon** W = 44, p = 8.364E-08 | | | | | |
|  | **Within Severe-acute group:**  **Two-sided Wilcoxon** W = 44, p = 2.30E-03  **ANCOVA adjusted for sex and age (using log10-transformation on biomarkers)**:  F(1,29) = 13.4579, p = 9.76E-04 | | | | | |
| **Figure 4B** | **GOSE Outcome (Favorable 5-8 vs. Unfavorable 1-4) differences in 3-step BD-tau** | | | | | |
|  | **Two-sided Wilcoxon** W = 51.5, p = 4.63E-07 | | | | | |
|  | **Within Severe-acute group:**  **Two-sided Wilcoxon** W = 51.5, p = 0.0081  **ANCOVA adjusted for sex and age (using log10-transformation on biomarkers)**:  F(1,28) = 1.852, p = 0.00239 | | | | | |

**Table 4.** Unadjusted model of performative prediction of Pitt-BD-tau in predicting TBI

| **Comparison** | **Measure** | **N** | **Thr** | **AUC** | **Sens** | **Spec** | **Accuracy** | **PPV** | **NPV** |
| --- | --- | --- | --- | --- | --- | --- | --- | --- | --- |
| Control vs Severe-acute | Quanterix BD-Tau | 40 | 2.81 | 1.000 | 0.969 | 1.000 | 0.975 | 1.000 | 0.889 |
| Control vs Severe-acute | 2STEP | 41 | 2.80 | 0.996 | 0.970 | 0.875 | 0.951 | 0.970 | 0.875 |
| Control vs Severe-acute | 3STEP | 40 | 3.31 | 0.998 | 0.969 | 0.875 | 0.950 | 0.969 | 0.875 |
| Chronic-mixed vs Severe-acute | Quanterix BD-Tau | 52 | 4.33 | 0.998 | 0.969 | 0.950 | 0.962 | 0.969 | 0.950 |
| Chronic-mixed vs Severe-acute | 2STEP | 54 | 3.13 | 0.996 | 0.970 | 0.952 | 0.963 | 0.970 | 0.952 |
| Chronic-mixed vs Severe-acute | 3STEP | 52 | 4.02 | 0.988 | 0.938 | 0.950 | 0.942 | 0.968 | 0.905 |

**Supplementary Figure**

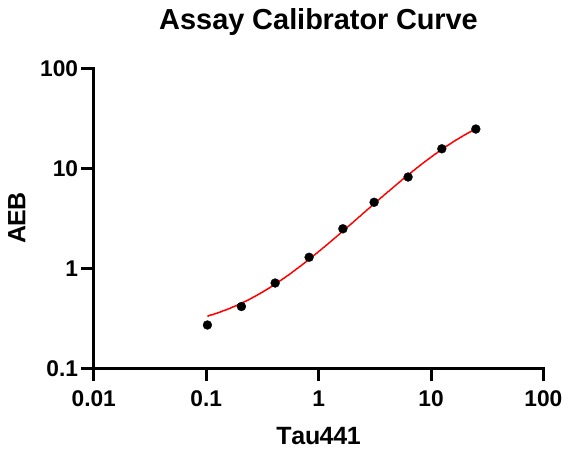

Figure 1: Four-parameter logistic (4PL) regression calibration curve on a log-log scale. The curve was generated with nine non-zero calibrators that are used with the Pitt-BD-tau assay.
